# Neurobehavioral Profiles and Clinical Consequences of MYT1L-Related Neurodevelopmental Disorder: Insights from the Brain Gene Registry

**DOI:** 10.64898/2025.12.01.25340510

**Authors:** Jorge L. Granadillo, Levi Kaster, Daleep Grewal, Allyson Schreiber, Anna M Abbacchi, Virginia Lanzotti, Brain Gene Registry Consortium, Aditi Gupta, Christina A. Gurnett, Kristen L. Kroll, Joseph D. Dougherty

## Abstract

*MYT1L*-Related Neurodevelopmental Disorder (*MYT1L*-NDD) is a rare autosomal dominant syndrome characterized by intellectual disability, global developmental delay, autism, and obesity. Despite growing recognition, prospective and systematic clinical phenotyping remains limited. Here, we analyzed data from 20 individuals with *MYT1L* variants enrolled in the Brain Gene Registry (BGR), a national platform integrating genetic, neurobehavioral, and clinical data. Neurobehavioral assessments were conducted using the Rapid Neurobehavioral Assessment Protocol (RNAP), including virtual adaptations of the Shipley, Developmental Profile-4, ASEBA Child Behavior Checklist, and Vineland-3. Diagnostic and medication data were extracted from structured electronic health records (EHR). *MYT1L* participants were compared to the broader BGR cohort. Fourteen participants had pathogenic or likely pathogenic *MYT1L* variants. Individuals with confirmed *MYT1L*-NDD exhibited significant impairments in nonverbal cognition and adaptive functioning, particularly in motor tasks, communication, and daily living skills. Behavioral assessments revealed elevated rates of ADHD symptoms, behavioral conduct problems, oppositional defiance, and anxiety. EHR data showed frequent diagnoses of developmental delays, speech and language disorders, gastrointestinal problems, and obesity. Medication data indicated common prescription of alpha-2 adrenergic agonists and laxatives. This study provides the first prospective, multi-source characterization of *MYT1L*-NDD. Findings expand our understanding of *MYT1L*-NDD phenotypes in a quantitative and systematic manner and underscore the value of registry-based approaches for understanding rare neurodevelopmental disorders. Larger, longitudinal studies are needed to further define *MYT1L*-NDD natural history and inform clinical care.

## Introduction

*MYT1L*-Related Neurodevelopmental disorder (*MYT1L*-NDD) is an autosomal dominant disease caused by pathogenic variants in *MYT1L*, a gene encoding a transcription factor essential for postmitotic neuronal maturation.^1^ The discovery and characterization of *MYT1L*-NDD has been driven by efforts of several teams over the last fifteen years. The first reported cases consisted of patients with intellectual disability (ID) who had a deletion at 2p25.3 spanning *MYT1L*.^2^ This was further supported by another study where five patients with loss of the paternal 2p25.3 chromosome involving *MYT1L* presented with various degrees of obesity, ID, and autism.^3^ Subsequently, De Rocker et. al (2015) determined that *MYT1L* disruption was the critical causative mechanism.^4^ This has been further confirmed by other researchers who have reported multiple individuals with *MYT1L* single nucleotide variants (SNVs; both truncating and non-truncating) with a similar presentation as those with 2p25.3 deletions.^5,6^ This underscores the substantial role that MYT1L has in neurodevelopment and its candidacy as the major pathogenic gene within this microdeletion.^5,7,8^

Investigations of *MYT1L*-NDD have begun to reveal a phenotypic profile characterized by global developmental delay, intellectual disabilities, autism, and obesity. To date, roughly 120 patients with *MYT1L* pathogenic variants– either SNVs or larger multigene deletions – have been reported.^9,10^ Coursimault et al. found that intellectual disability was present in most (70%) of the 62 patients with *MYT1L* SNVs (40 new patients and 22 previously reported cases), consistent with findings from previous research.^2,3,5,6,8,11^ Overweight and obesity rates hover around 50%.^2,3,6,8^ Among patients with *MYT1L* SNVs, autism has roughly a 30-40% prevalence.^5,6,8^

*MYT1L*-NDD is caused by three different types of *MYT1L* variants - missense variants, truncating variants, and 2p.25.3 microdeletions. Truncating variants in non-terminal exons are predicted to result in nonsense mediated decay and loss of function, suggesting haploinsufficiency as a major mechanism of *MYT1L*-NDD. However, few missense variants have been characterized, and genotype-phenotype correlations have not been characterized thus far. In a study on 62 patients with varying types of *MYT1L* variants, no statistically significant differences in the proportion of clinical features between these three groups were noted.^6^ By contrast, there are reports of individuals with partial duplications encompassing the 3’ end of the *MYT1L* gene that present with a different phenotype, characterized not by a childhood-onset neurodevelopmental disorder, but by schizophrenia in adulthood. Thus, this may reflect a different underlying mechanism, and the molecular consequences of these 3’ MYT1L partial duplications remain unstudied.^11^

Currently there is no specific treatment for *MYT1L*-NDD, and no evidence-based management guidelines are available. Medical care primarily focuses on ameliorating symptoms as they arise. Detailed phenotyping and application of standardized assessment tools are necessary steps to develop natural history studies and establish the foundations of clinical trial readiness. The Brain Gene Registry (BGR) is a trans-institutional data repository designed to study rare gene variants in putative brain genes by combining genetic data with existing electronic health record (EHR) data and neurobehavioral data collected from a novel, remotely administered phenotypic assessment battery – the Rapid Neurobehavioral Assessment Protocol (RNAP). The RNAP is a collection of instruments designed to provide a comprehensive neurobehavioral phenotype, which provides a snapshot of the individual’s cognitive, motor, psychiatric, and neurologic functioning. Over 300 brain genes are currently represented in the BGR, including *MYT1L*.^12^ Utilizing information gathered from the BGR can help establish practice and prescribing patterns for each unique monogenic disorder.

The present study leverages the BGR to provide a first-pass quantitative prospective assessment of patients with *MYT1L*-NDD, motivated by unanswered questions regarding the full phenotypic picture of patients with *MYT1L*-NDD and current treatment practices. Utilizing BGR data from enrolled MYT1L patients, we seek to 1) expand the sample size of reported patients with *MYT1L* variants to provide a clearer clinical description of *MYT1L*-NDD, 2) obtain a detailed and quantitative neurobehavioral characterization of an *MYT1L*-NDD cohort, and 3) test the utility of EHR data analysis for highlighting common diagnoses and prescription practices.

## Methods

### Recruitment and Data collection

Participants for this study were recruited to the Brain Gene Registry, as detailed in its initial publication.^12^ All participants, or their parents/legal guardians, gave informed consent.

The Washington University in St. Louis Institutional Review Board gave ethical approval for this work.

### Data commons platform – CIELO

BGR data sourced from GenomeConnect, RNAP, and EHR were stored in *CIELO –* Collaborative Informatics Environment for Learning on Health Outcomes *–* which allows for transfer and download of bundles of patient information, such as multi-scale, longitudinally-defined phenotypes, or basic information such as the recruiting institution or patient’s date of birth. We extracted de-identified patient bundles for all patients in the BGR, and analyzed these bundles with customized Python scripts (https://github.com/kasterlevi/MYT1L-NDD_Analysis), performing a focused analysis of patients with a variant in the MYT1L gene and using the remainder of the BGR as a comparison set.

### EHR data extraction and analysis

Patients receiving clinical care at specific Intellectual and Developmental Disabilities Research Centers (IDDRCs) had EHR data available for analysis. Notable elements of the EHR data available for analysis included medication orders, procedures, diagnoses, basic vitals, encounters, and clinical notes. The present study focused on medication orders amongst *MYT1L*-NDD patients to search for patterns of relevant and high-frequency drug usage. Medications were categorized by therapeutic categories and manually reviewed for clinical accuracy and relevance. We also report the frequency of different diagnostic categories in all BGR participants, and how these compare to our *MYT1L*-NDD subgroup. To evaluate patient diagnoses, we mapped all International Classification of Diseases (ICD) Codes to their corresponding Phecodes, which group ICD codes into clinically meaningful categories to support research and data analysis.^13–15^ ICD codes and phecodes were mapped according to Phecode Map 1.2 downloaded from PheWAS, which includes both ICD-9 and ICD-10 diagnoses.^16^

### Rapid Neurobehavioral Assessment Protocol (RNAP)

The BGR RNAP aggregates participant data from neurological, neuropsychiatric, and cognitive testing (full list described elsewhere ^12^). This was accomplished through direct assessments via a telehealth visit as well as with distributed surveys and questionnaires completed in REDCap (Research Electronic Data Capture) by either adult participants or caregivers of minors or adults unable to consent, as described.^12^ RNAP information collected on REDCap is securely transferred to CIELO in batches to be made available to CIELO users. In addition, we directly downloaded RNAP data from REDCap, to ensure access to recent assessments. Here, we analyzed the data from the Shipley, SRS-2, ASEBA-CBCL, and Vineland 3 instruments present in the RNAP, comparing *MYT1L*-NDD patients against the full BGR cohort and each instrument’s norms, where appropriate.

### GenomeConnect

Information on *MYT1L* variants was obtained from GenomeConnect, the ClinGen patient registry. Trained GenomeConnect personnel extracted this information from the clinical genetic testing reports submitted by the participants. Additionally, we manually reviewed each report to ensure accuracy, completeness, and to confirm variant classification, per ACMG guidelines.^17^

### Artificial Intelligence

WashU ChatGPT (https://gpt.wustl.edu) was utilized during the writing of this manuscript for assistance in improving spelling, grammar, redaction, and editing.

## Results

### Description of the Brain Gene Registry resource and participant demographics

We leveraged an existing framework for a gene-focused patient registry by partnering with the BGR (**Figure 1**), which curates genetic and medical health record data (where available) and prospectively assesses cognitive phenotypes with a remote phenotyping pipeline.

**Figure 1:**
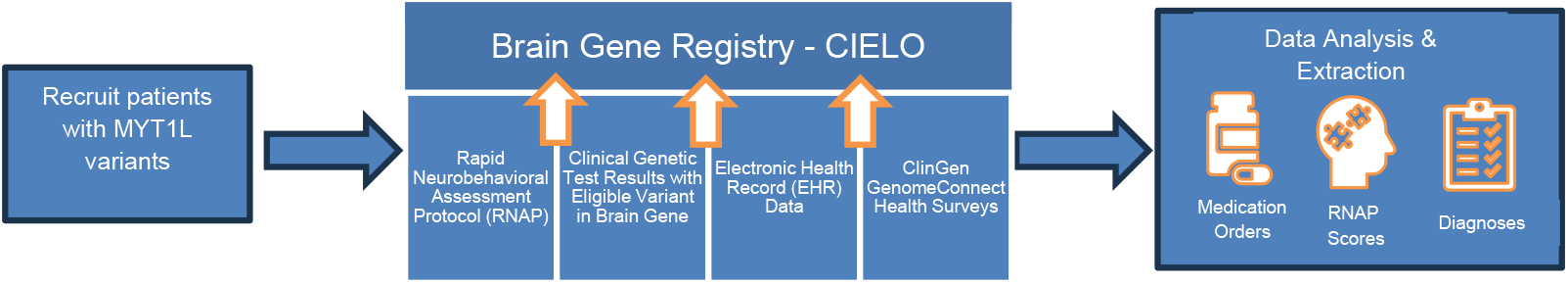
Summary of data extraction and analysis performed in the present study. Patients are first recruited based on having a known MYT1L gene variant. Rapid Neurobehavioral Assessment Protocol (RNAP) data – including cognitive ability, adaptive functioning, motor / sensory data, psychiatric symptoms, neurologic concerns, and physical features – are collected for each patient within the BGR. RNAP data is aggregated alongside EHR and ClinGen data to characterize the MYT1L cohort on the basis of medication orders, cognition scores, and specific diagnoses.

Based on data downloaded on 4/20/2024, there were 632 participants enrolled in the BGR, with 20 of these having a *MYT1L* variant and a completed CIELO profile (**Table 1**). Based on available genetic records, 14 (70%) participants had a *MYT1L* pathogenic or likely pathogenic variant and thus a confirmed diagnosis of *MYT1L*-NDD. Additionally, 6 patients (∼30%) reported variants of uncertain significance (VUS), which is slightly lower than the prevalence of VUS in the general BGR cohort (36%). Of these *MYT1L*-NDD participants, 8 (∼57%) had single nucleotide variants (SNV), and 6 (∼43%) had copy number variants (CNV). Missense variants constituted 28.6% of all *MYT1L*-NDD patients (4/14), 3 patients had SNVs that resulted in frameshift (21.4%), while one patient had a splicing variant. Reported pathogenic/likely pathogenic (P/LP) CNVs were expected to result in MYT1L haploinsufficiency through intragenic deletion (1), intragenic duplications (3), or multigene deletions (2). Ten (∼71%) of the P/LP variants were de novo, while 4 (∼29%) were of unknown inheritance. Inherited variants were only detected in participants with *MYT1L* VUSs. None of the MYT1L participants had a second P/LP variant. 57% (8/14) of patients with *MYT1L*-NDD were female, versus 36% male, and 7% unspecified. Interestingly, the general BGR cohort instead has a male predominance (55%), congruent with the previously reported higher general neurodevelopmental disorder prevalence in males^18^. Half of *MYT1L*-NDD patients identified as White, while this demographic information was unspecified for 21.4%. Other categories were represented by a single patient (Black/African American, other, more than one race). No *MYT1L*-NDD patient identified as Hispanic or Latino.

**Table 1:** Demographics and genetic variants of MYT1L participants enrolled in BGR and data commons platform.

| ID | Variant<sup>†</sup> | Protein | CNV nomenclature<sup>‡</sup> | Inheritance | Classification | Type | Sex | Race | Ethnicity |
| --- | --- | --- | --- | --- | --- | --- | --- | --- | --- |
| 1 | c.1585G>A | p.G529R | N/A | dn | Pathogenic | SNV missense | Male | Black or African American | NOT Hispanic or Latino |
| 2 | c.1585G>A | p.G529R | N/A | dn | Pathogenic | SNV missense | Female | White | NOT Hispanic or Latino |
| 3 | c.1633delG | p.V545Sfs*65 | N/A | dn | Pathogenic | SNV Deletion fs | Female | White | NOT Hispanic or Latino |
| 4 | c.2123dupG | p.S709Qfs*56 | N/A | dn | Pathogenic | SNV Duplication fs | Male | White | NOT Hispanic or Latino |
| 5 | c.2642+1G>A | N/A | N/A | Unk | Pathogenic | SNV splicing | Unk | Unk | Unk |
| 6 | 2.269Mb del | N/A | 2p25.3<br>(39896_2309039)x1 | Unk | Pathogenic | Multigene Deletion | Female | More Than One Race | Unk |
| 7 | 2.27Mb del | N/A | 2p25.3<br>(20341_2288046)x1 | dn | Pathogenic | Multigene Deletion | Female | Other | NOT Hispanic or Latino |
| 8 | Intragenic del. Exons 4-23 | N/A | 2p15.3<br>(1805171_2057923)x1 | dn | Pathogenic | Deletion intragenic | Male | Not Reported | Unk |
| 9 | c.1706G>A | p.R569Q | N/A | dn | Likely pathogenic | SNV missense | Male | Unk | Unk |
| 10 | c.178A>G | p.K60E | N/A | dn | Likely Pathogenic | SNV missense | Male | Unk | Unk |
| 11 | c.2726delG | p.G909Afs*31 | N/A | Unk | Likely Pathogenic | SNV Deletion fs | Female | White | NOT Hispanic or Latino |
| 12 | Intragenic dup. Exons 9-21 | N/A | 2p25.3<br>(1842905_1947121)x3 | dn | Likely pathogenic | Duplication Intragenic | Female | White | NOT Hispanic or Latino |
| 13 | Intragenic dup. Exons 6-21 | N/A | 2p25.3<br>(1929081_2257724)x3 | Unk | Likely Pathogenic | Duplication Intragenic | Female | White | NOT Hispanic or Latino |
| 14 | Intragenic dup. Exons 3-23 | N/A | Unk | dn | Likely Pathogenic | Duplication Intragenic | Female | White | NOT Hispanic or Latino |
| 15 | c.1184T>C | p.V395A | N/A | Mat | VUS | SNV missense | Male | Asian | Hispanic or Latino |
| 16 | c.1808G>T | p.R603L | N/A | Mat | VUS | SNV missense | Male | White | NOT Hispanic or Latino |
| 17 | c.1808G>T | p.R603L | N/A | Unk | VUS | SNV missense | Female | White | NOT Hispanic or Latino |
| 18 | c.2030A>G | p.D677G | N/A | Unk | VUS | SNV missense | Male | White | NOT Hispanic or Latino |
| 19 | 100.91Kb dup. Exons 21-25 of MYT1L. Exon 1 of PXDN | N/A | 2p25.3<br>(1742043_1842949)x3 | Unk | VUS | Duplication multiple genes | Male | Unk | Unk |
| 20 | 634K dup. Exons 1-9 of MYT1L and other genes | N/A | 2p25.3<br>(1245633_1979818)x3 | Mat | VUS | Duplication multiple genes | Male | White | NOT Hispanic or Latino |

Abbreviations: CNV, copy number variant; del, deletion; dup, duplication; mat, maternal; N/A, not applicable; pat, paternal; dn, *de novo*; SNV, single nucleotide variant; unk, unknown; VUS, variant of uncertain significance. <sup>†</sup>Reference Sequence NM\_001303052.2. <sup>‡</sup>Reference Genome GRCh37.

### Neurobehavioral Assessments

The results from cognitive tasks of the RNAP across MYT1L participants are summarized below. Given the diversity of age ranges (and thus differences in instruments used across patients), sample sizes were too low to warrant formal statistical analyses. However, individuals were coded by variant type and compared to the broader BGR sample, with general population norms and thresholds marked as well.

To assess nonverbal cognitive ability, the Shipley Block Patterns subtest was administered to participants able to actively participate in a video interview with the examiner, while the DP-4 (cognitive subscale) parent/caregiver checklist was utilized if participants were either too young or too severely affected to complete the Shipley (**Fig 2**). For MYT1L participants who completed the Shipley exam, scores were below the population mean for all participants (n=6), with 2 out of 3 individuals with confirmed *MYT1L*-NDD having scores at or below the threshold for intellectual disabilities (<70). Patients with MYT1L VUS (n=3) tended to have higher scores. On the other hand, the general BGR cohort had a broader distribution, with a mean score only slightly lower than the population mean, and the large majority of participants scoring above 70. For those MYT1L participants who completed the DP-4 instead (n=7), scores were overall lower, with the mean lower than the general population mean, and 67% (4/6) of patients with confirmed *MYT1L*-NDD scoring below the threshold for developmental delays (<70). Scores in the general BGR cohort had a broad distribution, but the mean was well below the general population mean and similar to the MYT1L subgroup.

**Figure 2:**
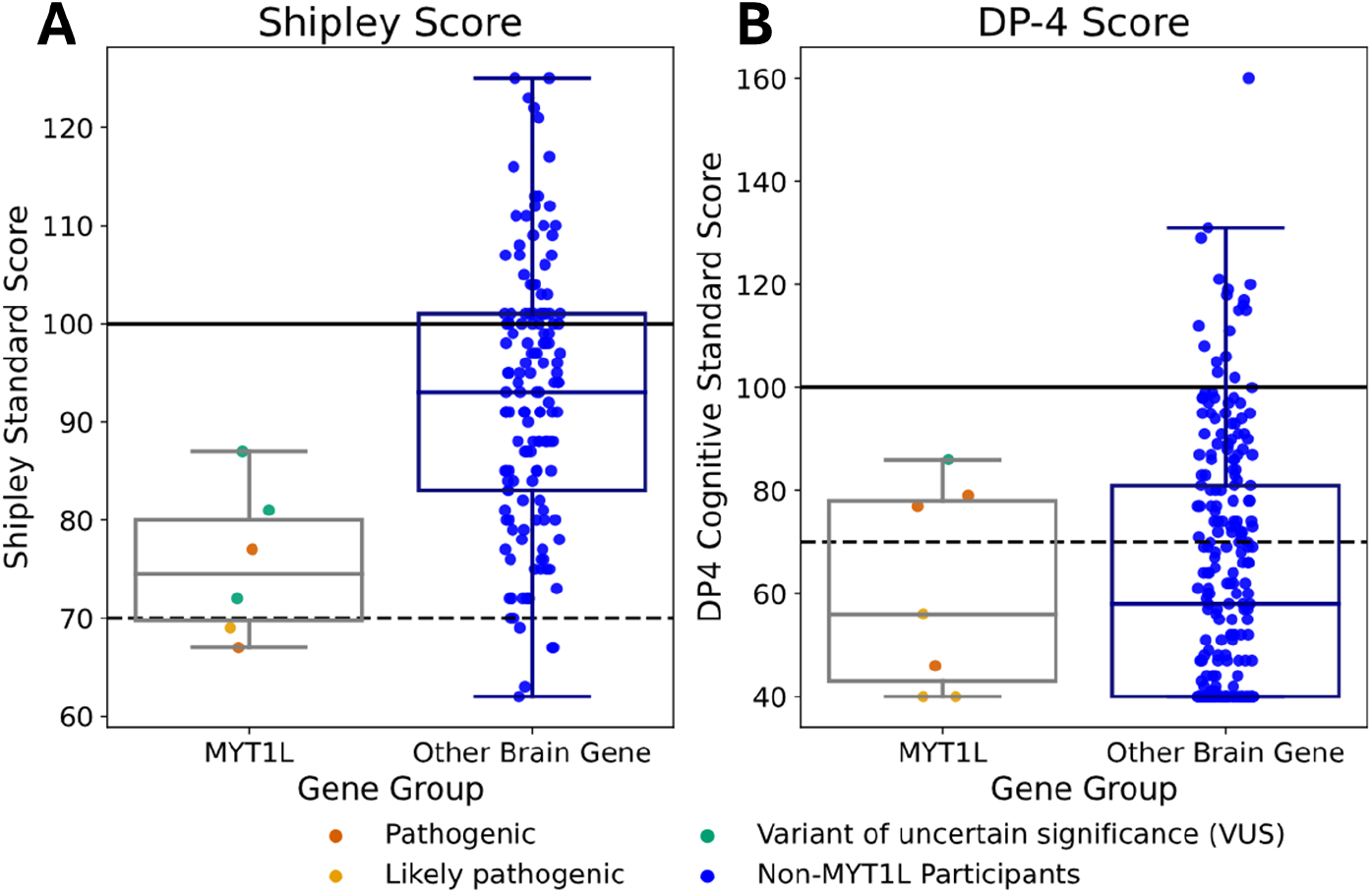
Cognitive scores for participants with MYT1L variants compared to all BGR participants A) Shipley assessment scores, MYT1L patients (N=6) versus patients with other gene variants, B) DP-4 assessment, MYT1L participants (N=7) versus patients with other gene variants. General population mean is marked with the solid line (100) and two standard deviations (dashed line; 70) is marked as a standard threshold for intellectual impairment.

The Child Behavior Checklist is the parent-completed component of the Achenbach System of Empirically Based Assessment (ASEBA), designed to assess emotional and/or behavior problems ^19,20^. Across all metrics of the ASEBA, T-scores lower than 65 indicate non-clinical symptoms, between 65-69 indicates at-risk behavioral issues, and 70 or higher indicates clear clinical symptoms. Sample sizes varied because each ASEBA subscale applies to a specific age range. Though most MYT1L participants were in the normal range **(Fig 3)**, a portion showed clinical symptoms for problems related to ADHD (27%, (33% excluding VUS)), conduct problems (29% (40% excluding VUS)), oppositional defiance (18% (22% excluding VUS)), anxiety (15% (20% excluding VUS)), and depression (8% (10% excluding VUS)). 50% were in the clinical range for autism, but this was based on only 4 individuals tested. No participant was in the clinical range for somatic problems. Avoidant personality problems or antisocial problems had small sample sizes (n=2) because these subcomponents only applied to participants age 18 and older. Participants with VUS generally performed better, with none of them at the clinical range.

**Figure 3:**
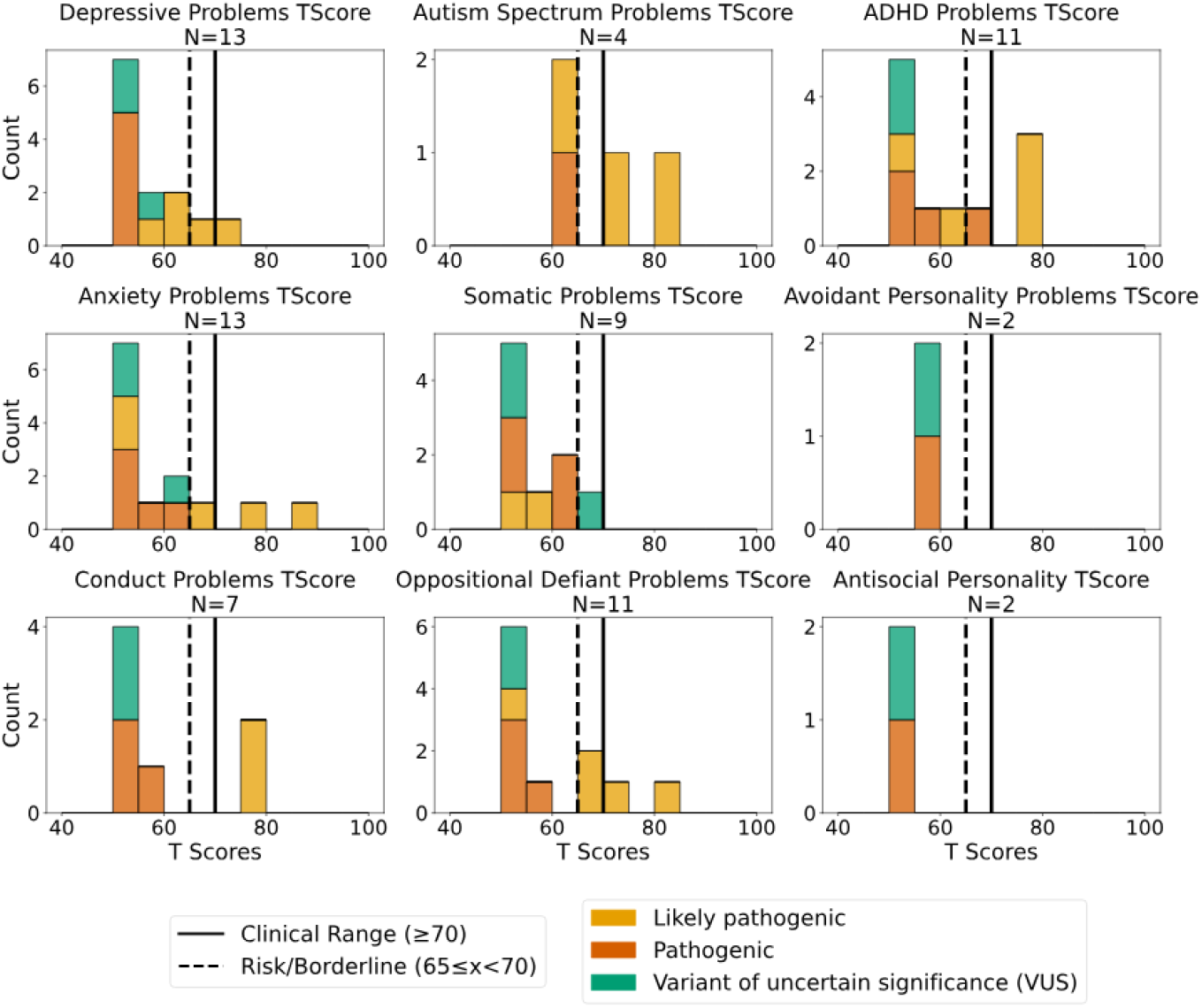
Distribution of ASEBA-CBCL behavior report T-scores for MYT1L patients within the registry. Vertical lines indicate thresholds for the at-risk or borderline range (65-69), the clinical range (≥70), and a normal score being <65.

The Vineland is a parent/caregiver-completed assessment designed to ascertain the extent of adaptive behavior in a child. Motor skills were assessed only in participants aged 9 years or younger. All MYT1L participants scored below the general population mean in both the composite score (ABC) and the subcomponents (**Fig 4**), with 54% (or 75% percent when VUSs are excluded) with ABC scores two standard deviations (SDs) below the population mean, showing a global decrease in adaptive behavior. The general BGR cohort scores were also below the general population mean, and scores mostly overlapped with those of MYT1L patients. This MYT1L cohort showed a higher prevalence of deficits in the motor skills subcomponent (71% < −2 SD (67% excluding VUS)), followed by communication (55% (64% excluding VUS)), Daily Living Skills (55% (63% excluding VUS)) and socialization (45% (50% excluding VUS)).

**Figure 4:**
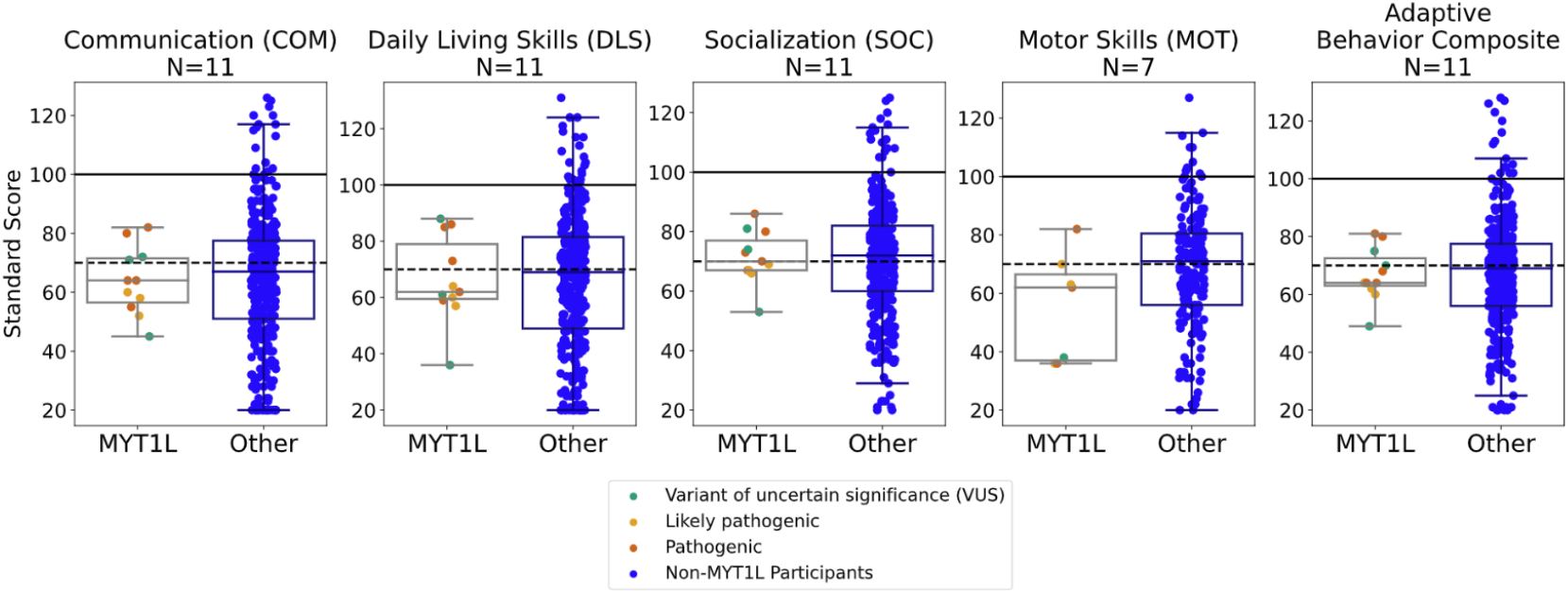
Distribution of Vineland-3 standard scores across 5 behavior categories. A) Distribution of scores observed in MYT1L cohort versus the general population (N=11/7). B) Distribution of scores in MYT1L cohort versus patients with other gene variants.

### Electronic Health Record (EHR) Data

Diagnosis data was obtained from the health record data contained within the BGR dataset for 9 MYT1L participants, 6 of whom had a P/LP *MYT1L* variant. These were categorized utilizing Phecodes **(Fig. 5)**.^15^

**Figure 5:**
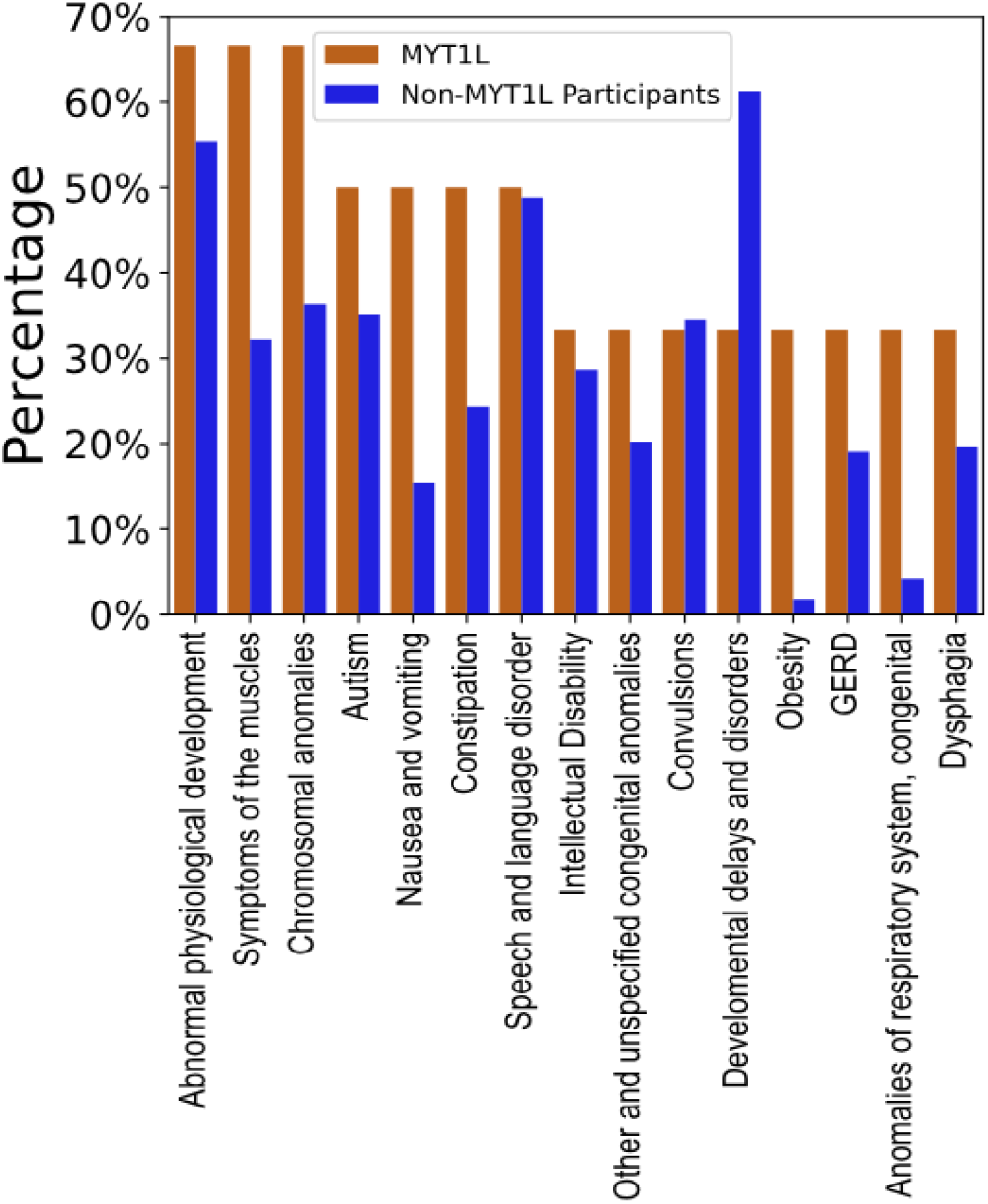
Distribution of the 15 most common Phecodes across available data for 6 MYT1L-NDD patients (VUS excluded), compared with the general BGR cohort. The Phecodes of “other dyspnea”, “pneumonia”, “fever of unknown origin”, “noninfectious gastroenteritis”, “overweight obesity and other hyperalimentation”, “decreased white blood cell count”, “delayed milestones”, “abnormality of gait”, and “amblyopia” were present at a 33% prevalence, but excluded from the figure for improved legibility and to decrease redundancy.

The most common Phecodes in patients with confirmed *MYT1L*-MDD were “abnormal physiological development (66% (4/6)), symptoms of the muscles (66% (4/6)), and chromosomal anomalies (66% (4/6)), followed by autism (50% (3/6)), gastrointestinal (GI)-related problems such as “nausea and vomiting” and “constipation” (50% each), and speech and language disorder (50%). Convulsions were found in a third of cases. Other diagnostic categories related to neurodevelopment were also highly prevalent, such as developmental delay (33% (2/6)), delayed milestones (33% (2/6)), and intellectual disability (33% (2/6)). This likely reflects the overlapping categories characteristic of Phecodes. Additional categories related to “Convulsions” were found at a lower prevalence, such as “epilepsy, recurrent seizures, convulsions”, “partial epilepsy”, and “epilepsy” at approximately 17% each (1/9). ADHD was found in 1 case (17%). Of note, a third of patients had a diagnosis of obesity, which is much higher than in the general BGR cohort (3%), and in line with previous reports.^6^ Other GI problems included GERD (2/6, (33%)), dysphagia (2/6, (33%)), and eating disorder (1/6 (17%)). Failure to thrive in childhood was found in one patient (17%). We analyzed the data for birth defects, although *MYT1L*-NDD is not typically associated with extra-neurological congenital anomalies. The Phecode “other and unspecified congenital anomalies” was found in 33% (2/6) *MYT1L*-NDD patients, higher than the general BGR cohort (19%); however, ICD-10 codes associated with this Phecode did not provide information about the specific anomalies. “Anomalies of the respiratory system, congenital” were found in two *MYT1L*-NDD patients (33%), associated with the ICD-10 codes for “laryngeal cleft” and “laryngeal stridor”. One patient had the Phecode “Esophageal atresia/tracheoesophageal fistula”, which is linked to the diagnostic code for “esophageal web”.

De-identified medication order data from CIELO was compiled to ascertain the most prescribed drugs for *MYT1L*-NDD patients **(Fig 6)**. Of the patients with *MYT1L* P/LP gene variants, 4 had medication order data from their EHR. These files contained the medication’s order data, start and end date, frequency, dosage, therapeutic class, and generic name.

**Figure 6:**
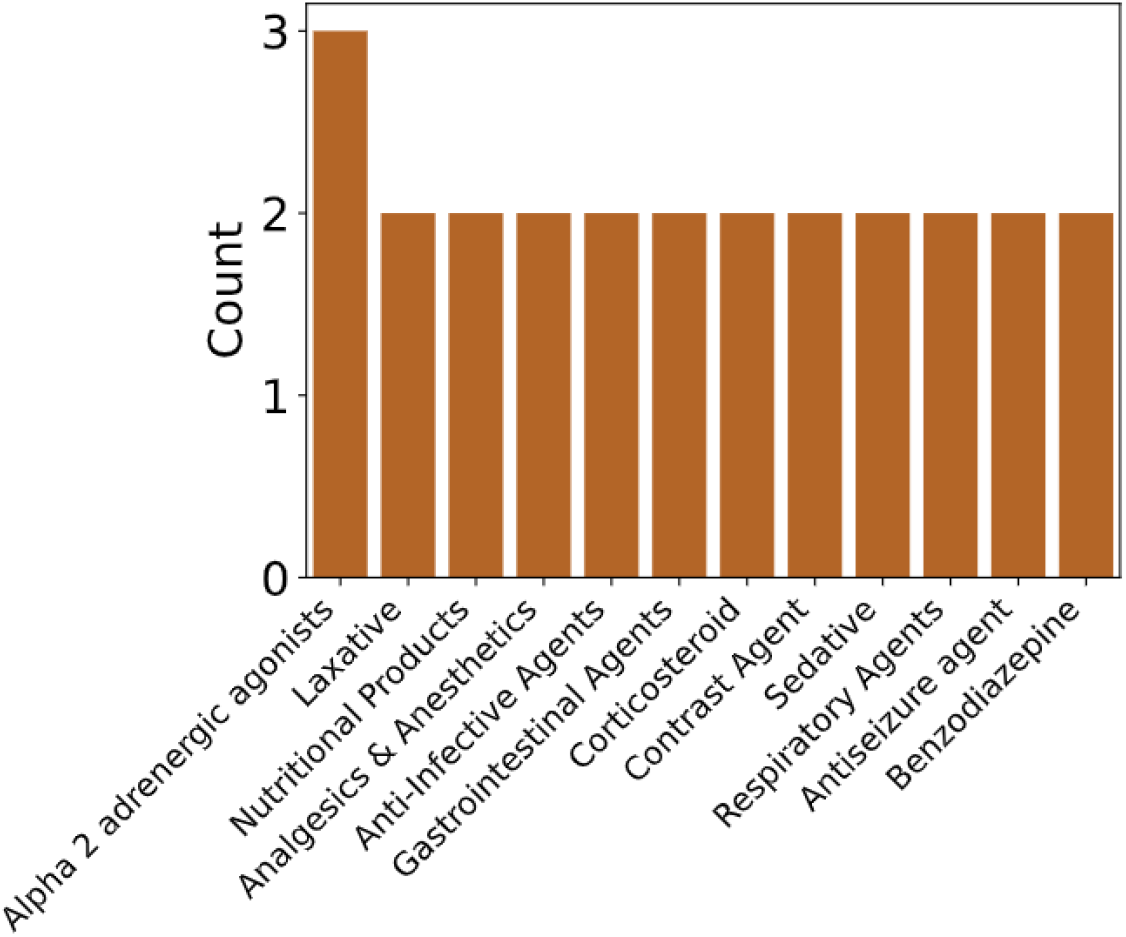
Distribution of medication orders by therapeutic category across available data for 4 MYT1L-NDD patients (VUS excluded).

Despite the small sample size, we analyzed how many individuals had been prescribed each drug at least once (**Fig 5**). The most frequently prescribed therapeutic class was alpha-2 adrenergic agonists (75% of individuals), namely guanfacine, which most likely reflects the high prevalence of ADHD in these patients, though does not correlate with the low prevalence of the ADHD Phecode in this group. This was followed by laxatives such as polyethylene glycol (PEG) (2/4), Other categories were highly prevalent, such as nutritional products, “analgesics & anesthetics”, and contrast agents. A deeper review of the data showed that this reflects inpatient orders related to medications and intravenous fluids in the setting of therapeutic and diagnostic procedures. Antiseizure medications were found in 2 out of 4 cases. Notably, lamotrigine has been shown to reverse behavioral phenotypes in MYT1L-mutant mice and rescue electrophysiological deficits in vitro.^21^ However, none of the study participants had been prescribed this medication.

## Discussion

This study represents a first attempt to quantitively and prospectively characterize *MYT1L*-NDD through the analysis of EHR data and standardized remote neurobehavioral research assessments through the BGR. Our findings confirm previous reports that *MYT1L* pathogenic variants are associated with intellectual disability, developmental delay, autism, and obesity. Consistent with prior literature, cognitive assessments such as the Shipley and DP-4 revealed significant impairment in nonverbal cognitive ability among *MYT1L*-NDD patients, both compared to population norms and other neurodevelopmental disorder cohorts within the BGR. Adaptive behavior deficits were also prominent, particularly in motor skills and communication domains, highlighting the broad impact of MYT1L variants on functional outcomes.

Behaviorally, ADHD symptoms were highly prevalent in our cohort, supported by both parent-reported ASEBA scores and medication prescribing patterns, notably the frequent use of guanfacine. This is in line with previous literature, where ADHD has been reported in 38% to 48% of *MYT1L*-NDD patients,^6,8,11^ warranting further investigation into targeted treatment strategies. Additionally, gastrointestinal issues such as constipation were common and more frequent than in the general BGR cohort, aligning with EHR data showing high rates of related diagnoses and laxative prescriptions. These findings underscore the importance of comprehensive clinical management addressing both neurological and systemic manifestations of *MYT1L*-NDD.

Our EHR analysis supports the frequent and prominent feature of obesity in *MYT1L*-NDD, with a prevalence much higher than in the general BGR cohort and consistent with previous reports. The pathophysiology of obesity in this population remains poorly understood. However, in our cohort, one patient (1/6, 17%) had a diagnosis of “compulsive eating patterns,” which may suggest that abnormal feeding behaviors may contribute to weight gain. Prior studies have also reported features such as hyperphagia, tachyphagia, and impaired satiety,^5–8^ indicating that increased intake and dysregulated appetite likely play a role in the development of obesity in *MYT1L*-NDD.

Extra-neurological anatomical anomalies have not been previously reported in patients with *MYT1L*-NDD. However, two patients in this cohort had a diagnosis of laryngeal cleft, and one patient had a diagnosis of esophageal web. Additional studies are needed to further confirm this finding and determine the role, if any, of *MYT1L* variants in GI and respiratory congenital anomalies.

The BGR intentionally includes participants with VUSs. Similarly, we have included MYT1L participants with VUSs in many of our analyses. This strategy serves two key purposes: it broadens the phenotypic spectrum captured by allowing for less restrictive recruitment, and it establishes a valuable, deeply characterized cohort for future variant curation and reclassification efforts. Among the 20 MYT1L participants, six carried VUSs, including a mother-son pair (participants 16 and 17). These individuals generally performed better on the RNAP assessments compared to those with confirmed *MYT1L*-NDD. Their variants were either missense or duplications involving multiple genes and the 5’ or 3’ end of *MYT1L*. Notably, patient 19 had a duplication involving the 3’-end of MYT1L. Preliminary evidence suggesting that these types of duplications may be associated with a different clinical entity characterized by increased risk for schizophrenia.^11^ Unfortunately, no EHR or RNAP data was available for this participant.

Our analysis of electronic health record data demonstrated the utility of leveraging real-world clinical information to identify common comorbidities and prescribing practices in rare neurodevelopmental disorders. However, limitations include the small sample size and incomplete data availability, which constrain statistical power, generalizability, and genotype-phenotype correlation analysis. Specifically, structured EHR data on medication orders and diagnostic codes were available for only 35% and 45% of participants, respectively. This may be related to the young age of some patients as well as the fragmentation of clinical care, which challenges the comprehensive capture of clinical data.

Future research should focus on expanding patient cohorts, longitudinal follow-up to delineate natural history, and functional studies to elucidate molecular mechanisms underlying phenotypic variability. Moreover, systematic evaluation of pharmacologic interventions, especially for ADHD and seizure management, is needed to inform evidence-based care guidelines. Our study focused on structured EHR data, but future work could leverage unstructured clinical notes using natural language models to gain deeper insights. The BGR platform offers a valuable resource for ongoing characterization and trial readiness in *MYT1L*-NDD and other monogenic neurodevelopmental syndromes. In summary, this study advances understanding of *MYT1L*-NDD by providing detailed neuropsychological profiles and highlighting key clinical features and treatment patterns. These insights lay groundwork for improved diagnosis, management, and therapeutic development for affected individuals and families.

## Author Contributions

Conceptualiztion and Methodology: JLG, JDD, CAG. Software, Data Curation: LK.

Formal Analysis: JG, LK.

Resources: JDD, AG, CAG, VL. Writing - Original Draft: JLG, DG, LK.

Writing - Review & Editing: JLG, LK, AS, VL, CAG, KLK, JDD Supervision: JLG, AG, CAG, JDD.

Funding acquisition: KLK, JDD.

## Data Availability Statement

The date that support the findings of this study are available from the corresponding author upon reasonable request.

## Acknowledgements

We would like to thank all BGR participants and their families and especially members of the MYT1L-NDD community, who contributed their time and effort to this study. This work was funded by P50 HD103525 (MPI: Dougherty, Marrus) and R01MH124808 (PI: Kroll). The Brain Gene Registry is supported by the IDDRC-CTSA Brain Gene Registry grant, U01TR002764, from the National Center for Advancing Translational Sciences (NCATS) of the National Institutes of Health (NIH), GenomeConnect is supported by U24HG006834 from the NIH National Human Genome Research Institute (NHGRI). We would also like to thank the Enrollment and RNAP team: Clarke, M^1^; Taylor, A^5^; Moreno Chaza, A^5^; German, R^3^; Cody, R^13^; Alagendran, K^13^; Savannah Erickson^13^; Nguyen, C^3^; Horn, I^10^; McNeil, D^9^; White, C^9^; Rusyniak, J^8^; Moradel Higareda, A^7^; Deppen, P^4^; Bican, A^11^; Rockouski, M^11^; Schneider, E^6^; Goodman, J^6^; Thompson, M^13^; Kinard, J^8^; Pandey, J^4^; Gomez^3^, V; Hut, A^3^; Grypp, D^10^

